# ATP-citrate lyase as a therapeutic target in chronic kidney disease: a Mendelian Randomization analysis

**DOI:** 10.1101/2021.10.07.21264710

**Authors:** Pedrum Mohammadi-Shemirani, Michael Chong, Nicolas Perrot, Marie Pigeyre, Gregory R. Steinberg, Guillaume Paré, Joan C. Krepinsky, Matthew B. Lanktree

## Abstract

**Background:** ATP-citrate lyase (ACLY) inhibition is a promising therapeutic target for dyslipidemia, atherosclerotic cardiovascular disease, non-alcoholic steatohepatitis, and metabolic syndrome. Genetic analysis of its role in chronic kidney disease (CKD) has not been performed.

**Methods:** We constructed a genetic instrument by selecting variants associated with *ACLY* expression level in the expression quantitative trait loci genetics consortium (eQTLGen) that includes blood samples from 31,684 participants. In a two-sample Mendelian randomization analysis, we then evaluated the effect of genetically predicted *ACLY* expression on risk of CKD, estimated glomerular filtration rate (eGFR), and microalbuminuria using the CKD Genetics consortium (CKDGen), United Kingdom biobank, and the Finnish Genetics consortium (FinnGen) totaling 66,396 CKD cases and 958,517 controls.

**Results:** *ACLY* is constitutively expressed in all cell types including in whole blood. The genetic instrument included 13 variants and explained 1.5% of variation in whole blood *ACLY* gene expression. A 34% reduction in genetically predicted *ACLY* expression was associated with a 0.04 mmol/L reduced low-density lipoprotein cholesterol (*P* = 3.4 × 10^−4^) and a 9% reduced risk of CKD (stage 3,4,5, dialysis or eGFR below 60 ml/min/1.73m^2^) (OR = 0.91, 95% C.I. 0.85-0.98, *P* = 0.008), but no association was observed with eGFR nor microalbuminuria.

**Conclusion:** Mendelian Randomization analysis provides cautious optimism regarding the possibility of ACLY as a therapeutic target for CKD.

## Background

Chronic kidney disease (CKD) remains an underappreciated cause of morbidity and mortality that is increasing in prevalence. CKD is a complex multifactorial syndrome and development of successful therapies has been challenging. Despite success with renin-angiotensin-aldosterone system (RAAS) blockade and sodium-glucose cotransporter 2 inhibitors (SGLT2i) there remains a significant need for development of novel therapeutic targets for CKD. Metabolic dysregulation, inflammation, and fibrosis are of great interest as they are key features of CKD not directly targeted by current treatments.

Mendelian randomization analysis is a valuable tool for evaluating investigational medical therapies and has rapidly grown in popularity.^1^ A genetic variant that alters the quantity or quality of a gene’s protein product can be used for a “natural” randomized trial. A single genetic variant often fails to explain enough variation in the quantity or activity of a protein product to evaluate its role in a disease, so either independent genetic variants are combined into scores at the individual-level or the effects of numerous independent variants are jointly assessed in a Mendelian randomization analysis.^2^ The genetic variants selected for inclusion in a Mendelian randomization analysis are referred to as the genetic instrument. Additional underlying assumptions of Mendelian randomization analysis need to be addressed with sensitivity analyses including: 1) the instrument explains a significant proportion of variation in the exposure of interest, 2) no association between the instrument and confounding factors, and 3) no association between the instrument and alternative causal pathways to the outcome except via the exposure.^2^ Quantitative trait loci (QTL) are genetic variants associated with continuous molecular traits, such as gene expression (eQTL), DNA methylation (meQTL), or protein concentration (pQTL).^3^ The eQTL genetics consortium (eQTLgen) performed a meta-analysis of genome-wide association studies of gene expression to successfully identify eQTL, creating a valuable resource for selecting genetic variants to be included in genetic instruments for Mendelian randomization analyses.^3^

ATP citrate lyase (ACLY) is a cytosolic enzyme that catalyzes the conversion of citrate into acetyl coenzyme A (acetyl-CoA), making it a critical intracellular switch to divert citrate between either oxidative phosphorylation in the citric acid cycle or to increase intracellular acetyl-CoA.^4,5^ Cytosolic acetyl-CoA can either lead to fatty acid and cholesterol synthesis or be moved into the nucleus where it modifies epigenetic DNA acetylation promoting transcription of inflammatory and fibrotic pathways such as nuclear factor kappa B and transforming growth factor beta (TFGβ).^6^ In both mice and humans inhibiting ACLY in the liver effectively reduces low-density lipoprotein (LDL) cholesterol,^4,5^ thus inhibition of ACLY was first evaluated as a treatment for dyslipidemia and cardiovascular disease. Mendelian randomization analysis of ACLY inhibition as a strategy for preventing cardiovascular disease was reported as supportive,^7^ but the variants included in their *ACLY* genetic instrument were only modestly associated with LDL cholesterol levels and coronary artery disease in additional populations calling the initial report into question.^8^ In clinical studies, bempedoic acid is a small molecule inhibitor of ACLY that safely lowered LDL cholesterol in multiple human clinical trials and a large outcome trial is ongoing.^9^

Lowering LDL cholesterol has proven ineffective for slowing progression of CKD in randomized controlled trials^10^ and Mendelian randomization studies,^11^ but findings in pre-clinical mouse models suggest ACLY inhibition could reduce inflammation and kidney fibrosis, making it an intriguing therapeutic target for CKD.^12,13^ Insulin resistance, obesity, non-alcoholic fatty liver disease, and proinflammatory cytokines such as interleukin-4 and interleukin-2, all of which are risk factors in CKD, lead to further increases in ACLY activity.^4,14,15^ Analyses of human genetic data regarding the importance of ACLY in kidney disease have yet to be reported. Using data from eQTLGen, we sought to create a genetic instrument for *ACLY* expression, evaluate the association between the instrument and LDL cholesterol to confirm its effect on ACLY activity, then assess the effect of the ACLY instrument on kidney phenotypes including prevalent CKD, estimated glomerular filtration rate (eGFR), and albumin-to-creatinine ratio (ACR) in population-level biobank data.

## Methods

### Creation of a genetic instrument for human ACLY expression

We first examined the tissue-specific expression profile of *ACLY* in humans in the Genotype-Tissue Expression Portal (GTEx).^16^ We identified genetic variants associated with *ACLY* expression in the eQTLGen Consortium dataset (https://www.eqtlgen.org/). eQTLGen performed eQTL genome-wide association studies (GWAS) of 16,989 genes in 31,684 participants from 37 cohorts and three gene expression platforms to identify variants associated with whole blood gene expression.^19^ We selected cis intronic genetic variants within 500kb of the *ACLY* gene that were associated with *ACLY* expression in whole blood (*P* < 0.05). Identified variants were then compared with the UK Biobank genotype dataset to avoid inclusion of eQTLGen variants that were unavailable in UK Biobank dataset. A prune-and-thresholding approach was utilized to identify independent genetic variants associated with *ACLY* expression. After identifying the most strongly associated variant, genetic variants in linkage disequilibrium with an r^2^ > 0.01 in 1000 genomes participants of European ancestry were pruned and discarded using the clump command in PLINK (http://zzz.bwh.harvard.edu/plink/).

### Study outcomes

eGFR was calculated from serum creatinine (eGFR_Crea_) or cystatin (eGFR_Cys_), age, and sex using the CKD-Epi equation.^20^ The primary outcome was the presence of CKD stage 3 or worse, defined as prevalent eGFR _Crea_ or eGFR Cys less than 60 ml/min/1.73m^2^. Secondary kidney outcomes included quantitative eGFR and the urinary ACR when available. Urinary albumin was determined in a spot urine sample in UK biobank; those with no detectable urinary albumin were set to the lower limit of detection and the albumin-to-creatinine ratio was log-transformed.

### Study Populations

After the genetic variants were selected, we first examined the strength of the ACLY expression genetic instrument based on the exposure r^2^ and F statistic using the two-sample MR package (https://rdrr.io/github/MRCIEU/TwoSampleMR/) in eQTLGen. Second, we assessed the cumulative effect of the genetic variants associated with ACLY expression by calculating an individual-level effect-weighted genetic score and testing its association with both LDL cholesterol and kidney traits in the European ancestry participants of the UK biobank (n = 343,648). Third, we evaluated the association in the 2019 Wuttke et al. public summary-level CKD association results of CKDGen (n = 522,093) accessed via https://ckdgen.imbi.uni-freiburg.de/.^21^ Finally, data from the Finnish genetics (FinnGen) consortium release 5 (n = 178,274) was accessed via www.finngen.fi. Data analyses in UK biobank were performed under UK Biobank application number 15255, while FinnGen and CKDGen data are publicly available. Notably there is no sample overlap between these three population-level datasets.

We also examined the effect of the *ACLY* expression instrument on eGFR in the summary-level results of the Stanzick et al. eGFR meta-analysis which includes the data from CKDGen and UK biobank (n = 1,201,930)^22^, and on the “rapid3” (greater than 3 ml/min/1.73m^2^ per year encompassing 34,874 cases and 107,090 controls) and “CKDi25” (25% or more decline in eGFRcrea or eGFRcys and crossing below 60 ml/min/1.73m^2^ encompassing 19, 901 cases and 175,244 controls) phenotypes as defined in the meta-analysis of Gorski et al.^23^ Note that these analyses include overlapping CKDGen samples and cannot be viewed as independent replications.

### Evaluation of rare genetics variants in ACLY by population sequencing

We next evaluated the prevalence of *ACLY* loss-of-function variants in the general population using the loss-of-function transcript effect estimator (LOFTEE) in gnomAD (https://gnomad.broadinstitute.org/)^17^. Genes that are essential for life contain less genetic variation in the general population.^17^ While the presence or absence of constraint against loss-of-function variants cannot nominate or exclude a gene as a drug target,^18^ they can be helpful to estimate the prevalence of those with a lifelong reduction in gene expression. Second, we looked for an excess burden of rare variants in those with coded “N18 Chronic Renal Failure” (n=9856) or “N17 Acute Renal Failure” (n=5015) out of 281,585 UK biobank participants in the gene-based association summary statistics phenome-wide association results (PheWAS) database Genebass (https://genebass.org/), and finally we tested for association of rare ACLY variants with lipid and kidney phenotypes in 173,688 UK biobank participants with available exome and quantitative phenotype data.

### Statistical analysis

Where individual-level genotype data was available (i.e. in the UK Biobank where every individual’s genotype is available), we constructed an *ACLY* expression genetic score for each individual. For each variant in the instrument, the number of expression-increasing alleles was coded as 0, 1, and 2 and multiplied by its normalized effect on *ACLY* expression to obtain the score. The specific genetic variants including in the genetic instrument and their weights are provided in Table S1. We then performed regression to evaluate the relationship between the *ACLY* instrument and kidney traits including adjustment for age, sex, and 20 principal components of ancestry.

Where only summary-level association results were available (i.e. the effect of each variant on the trait of interest in the complete sample in the remaining studies), we used the inverse variance weighted and weighted median methods regressing the genetic effect estimate of each variant on *ACLY* expression against the genetic effect estimate of each variant on the kidney trait. The effect estimates from the UK Biobank and summary-level CKDGen and FinnGen cohorts were then combined across all studies in a fixed-effect inverse-variance weighted Mendelian randomization analysis.

To evaluate the possibility of unmeasured directional pleiotropy we also performed Egger Mendelian randomization. As the effect of a variant on *ACLY* expression goes down to zero, its effect on the kidney trait should in theory also go to zero. Egger Mendelian randomization analysis allows for a non-zero Y-intercept in the Mendelian randomization regression line. Should a bias exist, the regression line does not go through the origin, and an effect on kidney traits appears to remain non-zero even as the effect of the variants on *ACLY* expression approaches zero. The Egger Mendelian randomization intercept test evaluates if the intercept is significantly different than zero. The primary limitation of Egger Mendelian randomization is a decrease in power, thus it is provided as a sensitivity analysis to evaluate for the possible presence of directional pleiotropy. In a second sensitivity analysis, we used Mendelian Randomization Pleiotropy RESidual Sum and Outlier (MR-PRESSO) to detect the presence of horizontal pleiotropy and remove outlier variants. All statistical analyses were conducted using R version 3.3.2 software.

## Results

### Development of the ACLY expression genetic instrument

Before developing a genetic instrument for *ACLY* expression, we verified that *ACLY* was expressed in whole blood, and compared the relative expression in whole blood to target organs including kidney, liver, and heart. *ACLY* is ubiquitously expressed with similar expression levels (transcripts per million reads) in kidney, liver, and whole blood (Figure 1). eQTLGen was developed by testing the association of genetic variants with expression of genes in whole blood. Thirteen non-coding variants within 500kb of *ACLY* were identified as independently associated with the quantity of *ACLY* expression (P < 0.05) and included in the genetic instrument (Table S1). Cumulatively, the instrument explained 1.5% of whole blood expression of *ACLY* (F = 32.9). In UK biobank, the ACLY expression genetic instrument was associated with LDL cholesterol (β = 0.04 mmol/L decrease per 34% relative reduction in *ACLY* expression, *P* = 3.4 × 10^−4^) and apolipoprotein B (β = 0.01 mmol/L decrease per 34% relative reduction in *ACLY* expression, P = 2.7 × 10^−5^) (Table S2). These observations are consistent with the ACLY eQTL genetic instrument impacting *ACLY* activity.

**Figure 1.**
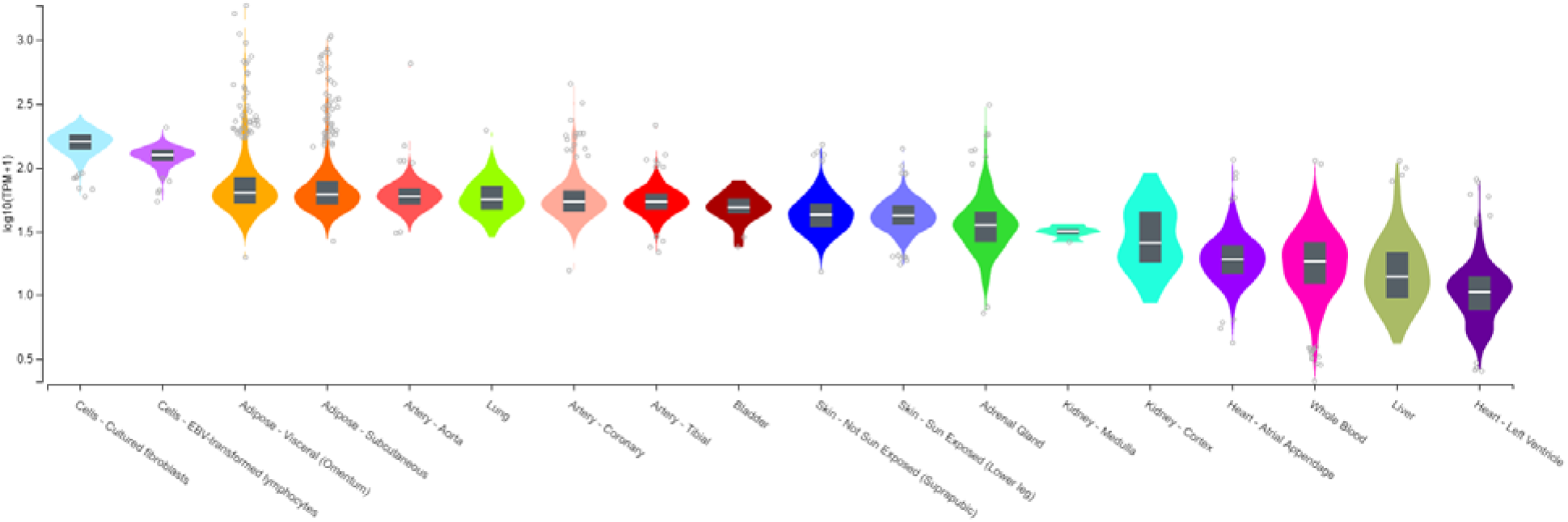
*ACLY* is ubiquitously expressed including in whole blood. TPM, transcripts per million.

### Association of ACLY expression genetic instrument with kidney outcomes

Meta-analysis of the primary CKD phenotype across UK Biobank, CKDGen, and FinnGen data including 66,396 CKD cases and 958,517 controls displayed a significant association between reduced ACLY expression and reduced risk of CKD (OR=0.91; 95% CI 0.85 to 0.98; *P* = 0.008, Figures 2 & S1). Examining the individual studies, the UK biobank white British ancestry participants included 22,291 cases with prevalent CKD stage 3, 4, or 5, or eGFR_Crea_ or eGFR _Cys_of less than 60 ml/min/1.73m^2^, and 321,357 participants without prevalent CKD. We observed an association between the *ACLY* genetic instrument and prevalent CKD (odds ratio (OR) = 0.91 per 34% relative reduction decrease in *ACLY* expression, 95% CI 0.82 - 1.00, *P* = 0.05). The *ACLY* expression instrument-CKD association was not statistically significant in 2-sample Mendelian randomization analysis of the summary level association results of either CKDGen or FinnGen, but the effect estimates were in the concordant direction with similar effect estimates as the UK biobank participants (OR = 0.91 per 34% relative reduction in expression score, 95% CI 0.81-1.02, *P* = 0.09 in CKDGen and OR = 0.93, 95% CI 0.74 – 1.18, P = 0.55 in FinnGen, Figure S2). For all MR analyses the Egger regression found no significant evidence for a non-zero intercept. MR-PRESSO did not identify evidence of horizontal pleiotropy or outlier variants.

**Figure 2.**
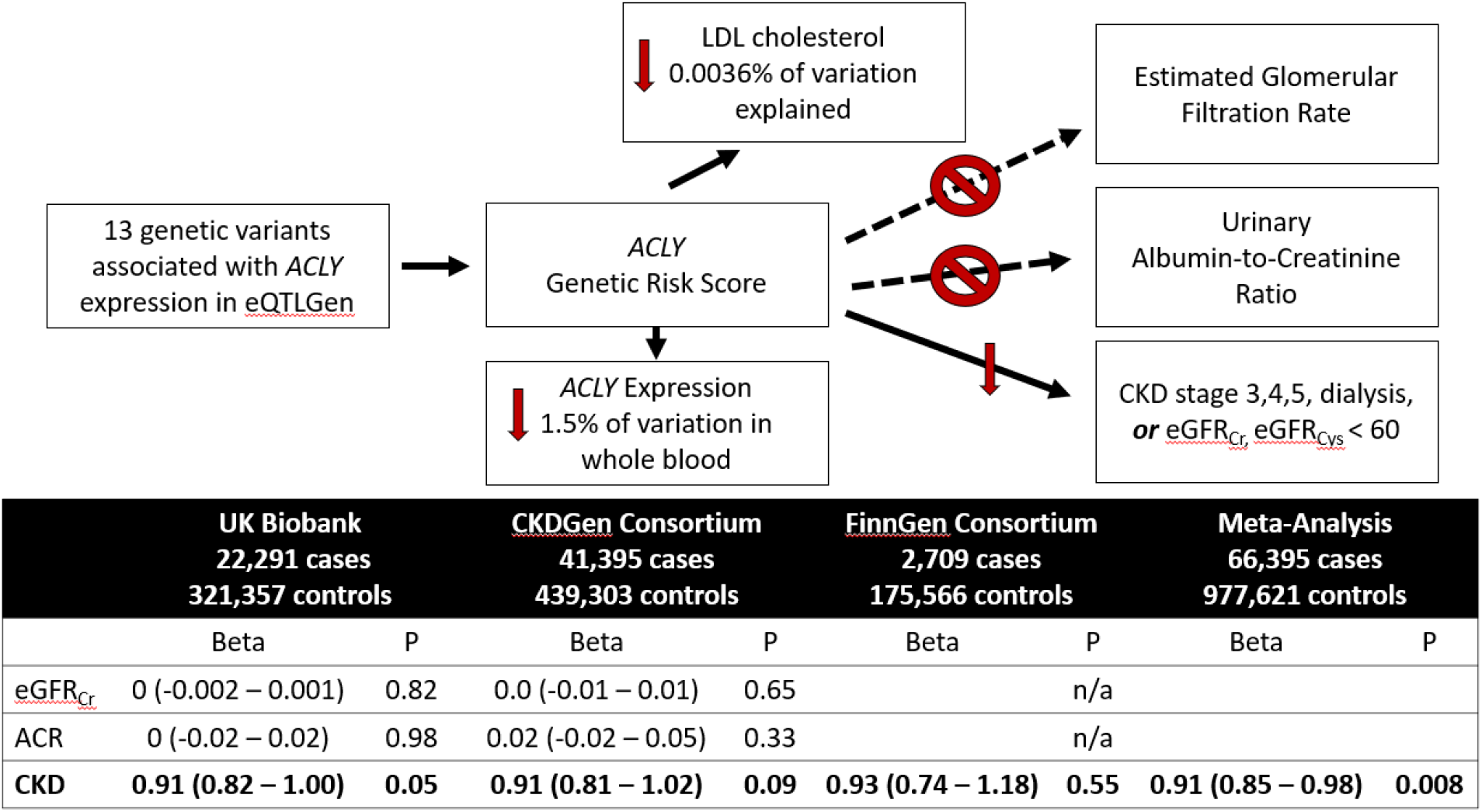
Mendelian randomization analysis supports reduced *ACLY* expression creates reduced risk of CKD without impacting eGFR. Beta given as trait change or odds ratio per 34% reduction in genetically determined *ACLY* expression.

In both individual-level Mendelian randomization analysis within the UK biobank white British ancestry participants and summary-level Mendelian randomization analysis with CKDGen our analysis displayed no association between the *ACLY* expression instrument and either eGFR_Crea_, eGFR_Cys,_ nor ACR (*P* > 0.3). Two eGFR meta-analyses published in 2021 included both CKDGen and UK Biobank populations. Using the Stanzick et al. eGFR meta-analysis results, there was again no observed Mendelian randomization association between the *ACLY* expression genetic instrument and either eGFR phenotype (eGFRcrea *P* = 0.83 and eGFRcys *P* = 0.73, Table S3).^22^ Using the Gorski et al. meta-analysis of rapid kidney function decline results, there was a nominal Mendelian randomization association of the ACLY expression genetic instrument with the dichotomous “Rapid3” greater than 3 ml/min/1.73m^2^ per year phenotype (OR=0.88; 95% CI 0.78 to 1.00; *P* = 0.05, Table S3) with the weighted median model, while the inverse variance weighted model was just above the significance threshold (OR=0.88; 95% CI 0.76 to 1.01; *P* = 0.07, Table S3). There was no association seen with the dichotomous “CKDi25” >25% drop across the 60 ml/min/1.73m^2^ threshold (*P* > 0.5).^23^

### Population-level analysis of rare genetic variants in ACLY

No monogenic phenotype has been reported to be associated with rare pathogenic variants in ACLY. Exome sequencing of control populations reported in gnomAD identified 15 loss-of-function ACLY genetic variants, and zero homozygous human *ACLY* “knockouts” in 141,456 sequenced exomes. This corresponds to a carrier prevalence of heterozygous *ACLY* loss-of-function mutations of 1 in 1,221 people in the general population. There were 64% fewer missense variants (95% confidence interval [CI] 59% to 70%) and 24% fewer loss-of-function mutations (95% CI 16% to 38%) compared to the expectation based on the distribution of rare variants in all genes in the genome (based on the gnomAD *observed / expected score*). In the 281,585 UK biobank participants in Genebass, there was no excess burden of loss-of-function variants in participants with “N18 Chronic Renal Failure” *(P* = 0.11) or “N17 Acute Renal Failure” (*P* = 0.63), nor with any of the other 1756 phenotypes in the GeneBass PheWAS below a Bonferroni-corrected significance (P<5×10^−5^). In the 173,688 participants of UK biobank with individual-level exome and quantitative trait data available to our group, there were 51 carriers with heterozygous predicted loss-of-function variants and there was no association with the positive control, LDL cholesterol (*P* = 0.91) (Table S4).

## Discussion

Using Mendelian randomization analysis of cis genetic variants associated with *ACLY* expression and population-scale genomic studies of kidney phenotypes, we found evidence that genetically reduced *ACLY* expression was associated with reduced risk of CKD despite having no association with eGFR nor urinary ACR. This is an apparent contradiction given that the presence of CKD is defined by a reduced eGFR and increased ACR. However, a genetic variant that alters kidney fibrosis or inflammation could increase risk of CKD development without a measurable effect on eGFR, especially when examining populations with largely normal kidney function. Similar relationships have been observed in other continuous complex traits with discrete diagnostic thresholds, such as the differing genetic architectures for fasting glucose and risk of type 2 diabetes, even though the diagnosis of type 2 diabetes can be made using fasting glucose.^24^

Mendelian randomization analysis of ACLY inhibition was initially reported with an odds ratio of 0.82 for major cardiovascular events per 10 mg per deciliter (0.26 mmol/L) decrease in LDL cholesterol by *ACLY* score.^7^ The analysis was criticized as many variants included in their *ACLY* genetic instrument were selected for their association with LDL cholesterol not ACLY activity, and were modestly associated with LDL-cholesterol levels or coronary artery disease in additional populations.^8^ We used a superior approach by selecting variants associated with *ACLY* expression, as opposed to selecting variants based on their association with LDL cholesterol. None of the variants in the current study overlap with those used in the Ference *et al.* study.

The largest clinical trial of pharmacological inhibition of ACLY was the Cholesterol Lowering via bempedoic acid, an ACL-Inhibiting Regimen (CLEAR) Harmony trial that enrolled 2230 people with high vascular risk and LDL cholesterol greater than 70 mg per deciliter despite maximally tolerated statin therapy. Daily bempedoic acid for 52 weeks lowered mean LDL cholesterol levels by 19.2 mg per deciliter (0.50 mmol/L, 18.1%) and did not increase the incidence of adverse or serious adverse events compared to placebo^25^. Our *ACLY* expression instrument tested here was associated with only a 0.04 mmol/L reduction in LDL cholesterol concentrations in UK Biobank, so bempedoic acid could be expected to have larger effects on CKD reduction than the 9% reduction in odds ratio we observed. There was a statistically significant but clinically irrelevant difference in serum creatinine levels in CLEAR Harmony participants who did not have kidney disease, with a 0.02 ± 0.13 mg/dl increase in the bempedoic acid arm compared to a 0.02 ± 0.12 mg/dl decrease in the placebo arm, which was postulated to be related to kidney transporter competition between creatinine and the drug.^25^ The CLEAR Harmony trial was of insufficient duration to evaluate cardiovascular outcomes, but we await the results of the CLEAR Outcomes trial that has finished recruitment of 14,000 statin-intolerant patients in 2019. Although not a pre-specified endpoint of the CLEAR Outcomes, retrospective analysis of differences in the development of CKD between those treated with bempedoic acid or placebo are expected to be available.

A potentially important difference between the Mendelian randomization analysis of *ACLY* expression and clinical trials with bempedoic acid could be related to the systemic effects of *ACLY* expression in the Mendelian randomization analysis compared to the tissue-targeted effects of bempedoic acid. *ACLY* is ubiquitously expressed and the Mendelian randomization analysis assumes reductions in whole-blood *ACLY* are reflected in all tissues to a similar degree. This contrasts with bempedoic acid which is only expected to inhibit ACLY in a small number of cell types and tissues. The selectivity of bempedoic acid for inhibiting ACLY has been shown to be dependent on the conversion of the drug to bempedoic acid-CoA by the long-chain acyl-CoA synthetase-1 (ACSVL1/SLC27A2).^5^ *SLC27A2* is minimally expressed in most cell types creating tissue specificity for bempedoic acid. However, GTEx indicates kidney cortex is second only to the liver for *SLC27A2* expression suggesting bempedoic acid would also inhibit ACLY in the kidney (Figure S3).

As ACLY catalyzes the conversion of citrate into acetyl-CoA, it is a reasonable hypothesis that inhibition of ACLY would lead to an increase in intracellular citrate concentrations. Urinary citrate is inversely correlated with glomerular filtration rate and acid retention.^26^ Low urinary citrate has also been recognized in autosomal dominant polycystic kidney disease.^27^ Whether either pharmacologic inhibition of ACLY or reduced *ACLY* expression would subsequently alter urinary citrate concentrations and whether urinary citrate could potentially be studied as a biomarker of kidney ACLY inhibition is yet unknown.

Strengths of this work include using the eQTLGen consortium to select genetic variants associated with *ACLY* expression and large population-based cohorts for the two sample Mendelian randomization analysis. We tested variants associated with *ACLY* expression, which is likely an imperfect proxy for ACLY activity, but testing for association with LDL cholesterol concentration serves as a positive control. *ACLY* is ubiquitously expressed which does lead to a lack of specificity if it is to be used as a therapeutic target, but bempedoic acid’s specificity for tissues with *SLC25A2* expression and its safety have already been assessed. All Mendelian randomization analyses are limited by the possibility of unmeasured pleiotropy, where the outcome is impacted by the genetic variant through a pathway different than the exposure, but the possibility of pleiotropy is greatly reduced using a 13-variant instrument nearby the encoding gene as opposed to hundreds of variants throughout the genome. Notably, the observed effect size, a 9% reduced risk of CKD for a 34% reduction in *ACLY* expression is small and required large sample sizes to identify, but bempedoic acid treatment had larger effects on LDL cholesterol and thus could be hypothesized to have a larger absolute effect on CKD prevention than our genetic instrument.

In conclusion, Mendelian randomization analyses supports that genetically reduced *ACLY* expression leads to a modestly reduced risk of CKD, despite having no measurable effects on eGFR or microalbuminuria.

## Data Availability

All data produced in the present study are available from public resources and results are presented in the manuscript. Any further data is available upon reasonable request to the authors.

https://genebass.org/

https://gnomad.broadinstitute.org/

https://ckdgen.imbi.uni-freiburg.de/

https://www.finngen.fi

https://www.eqtlgen.org/

https://biobank.ndph.ox.ac.uk/showcase/

## Author contributions

MBL, JK, and GRS conceived of the study. PM and MC performed data analysis. MBL drafted the manuscript. All authors contributed to study design, data interpretation, critical revisions, and provided approval of the final draft.

## Acknowledgements

The authors thank the participants and investigators of the contributing consortiums including eQTLGen, UK biobank, CKDGen, and FinnGen. MC acknowledges support by a CIHR Frederick Banting and Charles Best Canada Graduate Scholarships Doctoral Award. GRS acknowledges the support of a Diabetes Canada Investigator Award (DI-5-17-5302-GS), a Canadian Institutes of Health Research Foundation Grant (201709FDN-CEBA-116200), a Tier 1 Canada Research Chair in Metabolic Diseases and a J. Bruce Duncan Endowed Chair in Metabolic Diseases. JCK acknowledges support from the Canadian Institutes of Health Research (PJT-162411) and Kidney Foundation of Canada. JCK and MBL acknowledge support of the Research Institute at St. Joe’s Hamilton for nephrology research. MBL is a new investigator in the KRESCENT (Kidney Research Scientist Core Education and National Training) program funded by the Canadian Institutes of Health Research, Kidney Foundation of Canada and the Canadian Society of Nephrology.

## Supplemental material table of contents

**Table S1**. Markers included in *ACLY* expression genetic instrument from eQTLGen.

**Table S2**. Mendelian Randomization of *ACLY* expression instrument with lipid and kidney traits in the UK Biobank.

**Table S3**. Two-sample summary-level Mendelian randomization analyses in CKDGen and FinnGen.

**Table S4**. Rare variant association results for UK biobank.

**Figure S1**. Two sample Mendelian randomization plots of the instrument effect on *ACLY* expression and risk of CKD (A), Urine ACR (B), and eGFR (C) in CKDGen.

**Figure S2**. Two sample Mendelian randomization plots of the instrument effect on *ACLY* expression and risk of CKD in FinnGen.

**Figure S3**. GTEx tissue-specific expression of *SLC27A2* which is required for the activation of bempedoic acid.

