## Supplementary Figures for "ATP-citrate lyase as a therapeutic target in chronic kidney disease: a Mendelian Randomization analysis"

### Slide 1
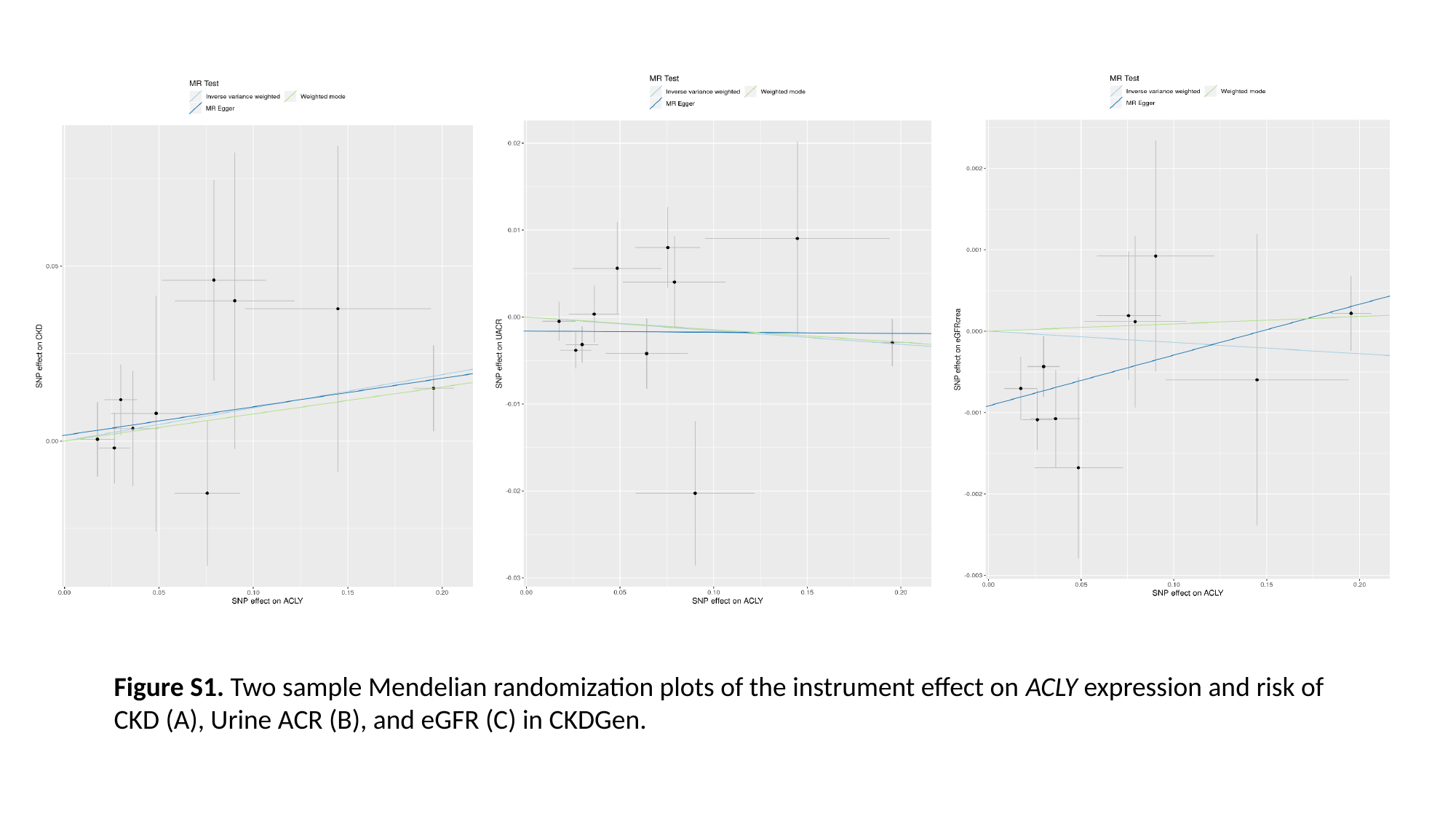

Figure S1. Two sample Mendelian randomization plots of the instrument effect on ACLY expression and risk of CKD (A), Urine ACR (B), and eGFR (C) in CKDGen.

### Slide 2
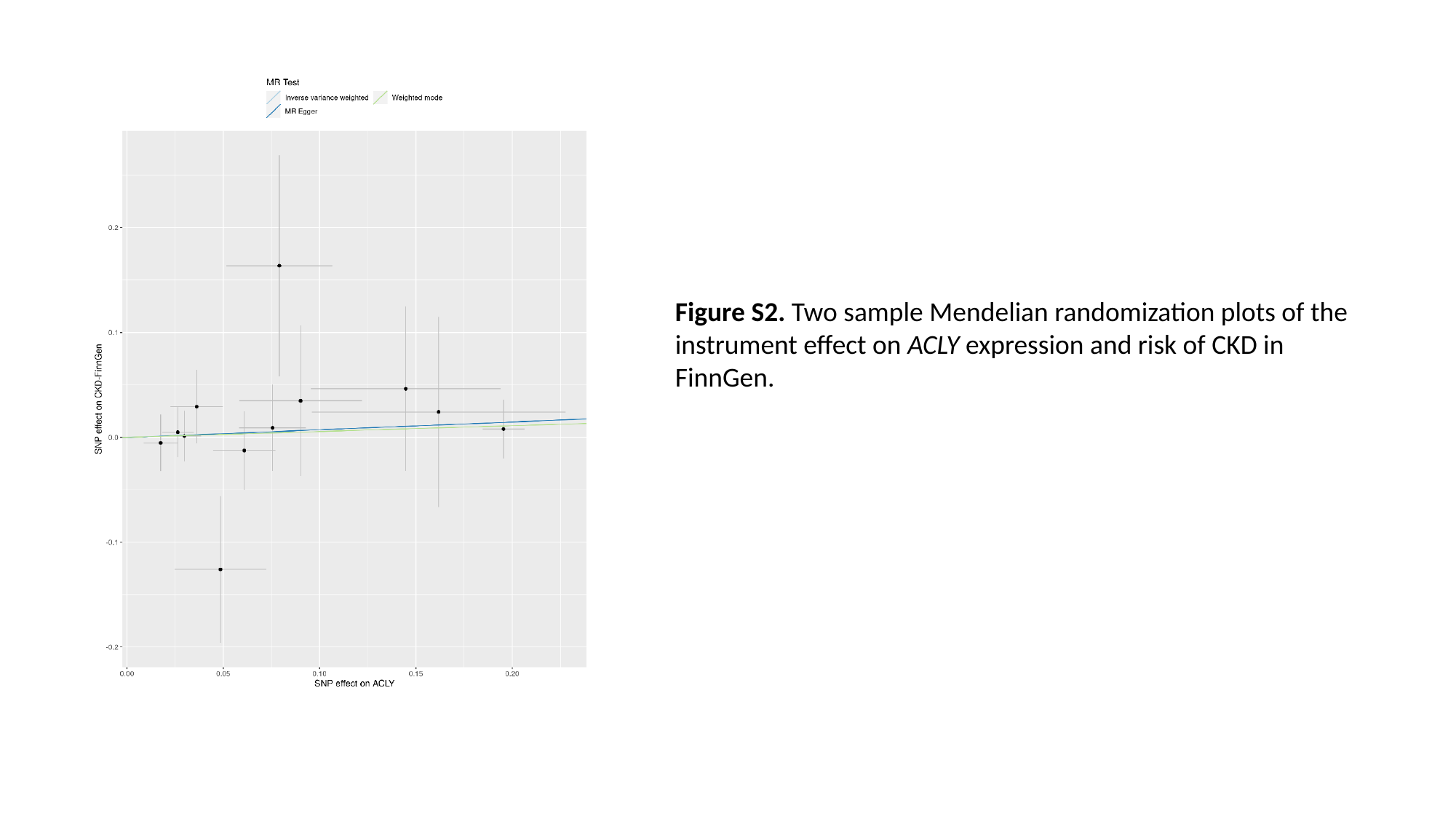

Figure S2. Two sample Mendelian randomization plots of the instrument effect on ACLY expression and risk of CKD in FinnGen.

### Slide 3
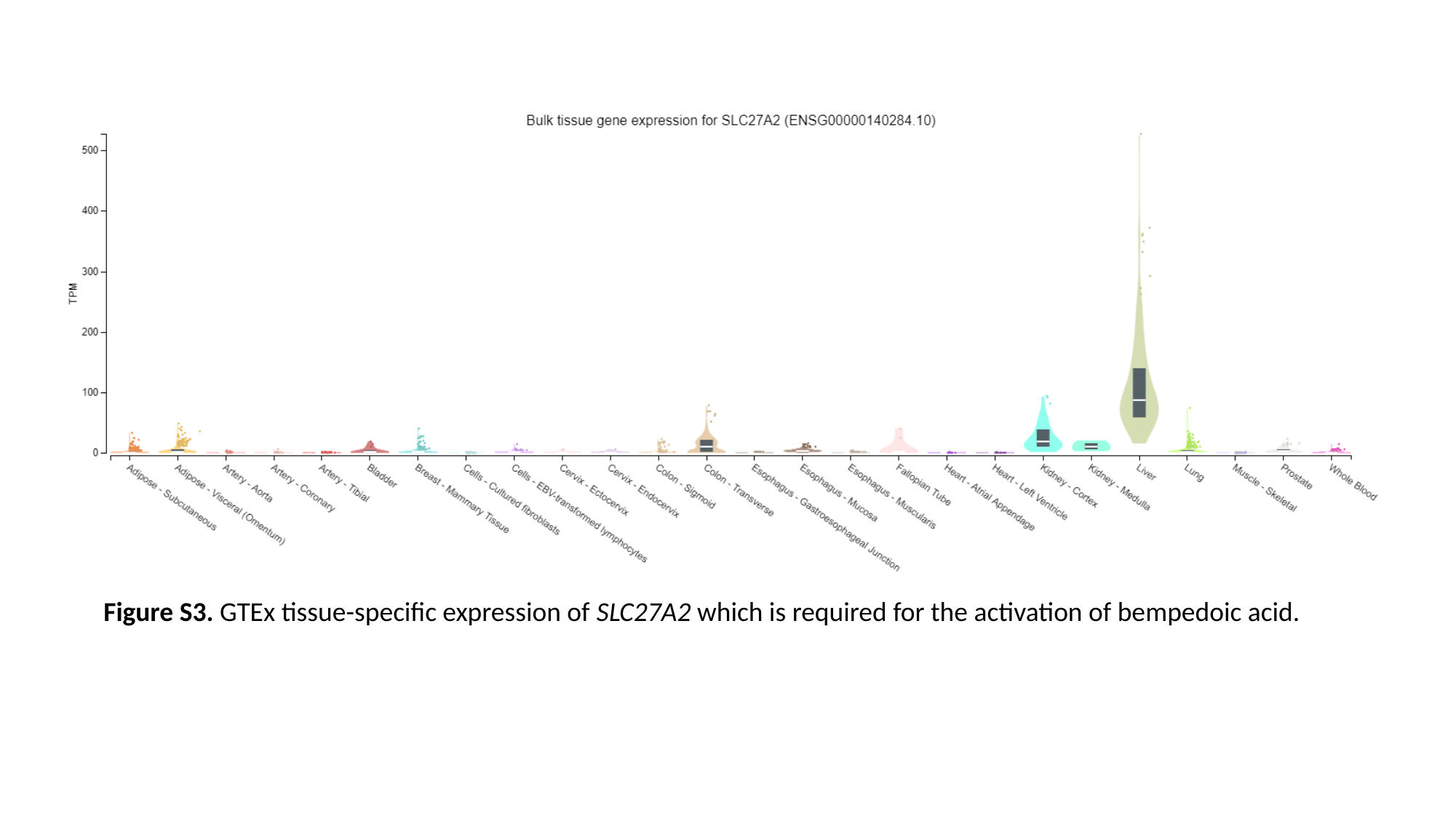

Figure S3. GTEx tissue-specific expression of SLC27A2 which is required for the activation of bempedoic acid.
